# Predicted domination of variant Delta of SARS-CoV-2 before Tokyo Olympic games, Japan

**DOI:** 10.1101/2021.06.12.21258835

**Authors:** Kimihito Ito, Chayada Piantham, Hiroshi Nishiura

## Abstract

Using numbers of SARS-CoV-2 variants detected in Japan, the relative instantaneous reproduction numbers of the R.1, Alpha, and Delta variants with respect to other strains circulating in Japan were estimated at 1.245, 1.437, and 1.948, respectively. The numbers can vary within 1.190–1.319 for R.1, 1.335–1.580 for Alpha, and 1.703–2.30 for Delta depending on the assumed serial interval distributions. The frequency of the Delta is expected to take over the Alpha in Japan around July 12, 2021.

---

Severe acute respiratory syndrome coronavirus 2 (SARS-CoV-2), the causative agent of COVID-19, has undergone adaptive evolution since its emergence in the human population in 2019. On May 31, 2021, the World Health Organization (WHO) has designated four variants SARS-CoV-2 as Variants of Concerns (VOCs)—Alpha, a variant lineage first reported in the United Kingdom; Beta, those in South Africa; Gamma, those in Brazil; Delta, those in India (World Health Organization, 2021). Volz et al. estimated 50–100% increase in *R* of the Alpha variant also using data from England (Volz et al., 2021). In our previous paper, we estimated the selective advantage of the Alpha variant in the England to be 26–45% higher than pre-existing strains using GISAID sequence data collected in England (Piantham, Linton, Nishiura, & Ito, 2021). These analyses use models that assume two circulating variants. As of June 9, 2021, the state of emergency has been declared in ten prefectures in Japan—Tokyo, Kyoto, Osaka, Hyogo, Aichi, Fukuoka, Hokkaido, Okayama, Hiroshima, and Okinawa. Due to the high transmissibility of VOCs, the replacement from local strains to Alpha and Delta variants is progressing, posing a serious public health threat in Japan. In such a situation where multiple variants are co-circulating, a model describing the adaptive evolution among multiple variants is required. Using numbers of SARS-CoV-2 variants found in the GISAID database (Shu & McCauley, 2017) and those detected by PCR in Tokyo, we estimate the relative instantaneous reproduction numbers of the R.1, Alpha, and Delta variants with respect to other strains circulating in Japan. We also show the expected temporal changes in variant frequencies of SARS-CoV-2 in Japan until early August 2021.

## Model of Advantageous Selection Among Multiple Variants

Suppose that we have a large population of viruses consisting of strains of genotypes *a, A*_1_, …, *A*_*n*_, of which frequencies in the viral population at a calendar time *t* are 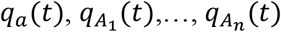, respectively. Suppose also that genotype *a* is a wildtype virus that was circulating at the beginning of the target period of analysis and that genotype *A*_1_,…, *A*_*n*_ are mutants that introduced to the population at time *t*_1_,…, *t*_*n*_, respectively.

We assume that viruses of genotype *A*_1_,…, *A*_*n*_ generate 1 + *s*_1_,…, 1 + *s*_*n*_ times as many secondary transmissions as those of genotype *a*, respectively. The instantaneous reproduction number is defined as the average number of people someone infected at time *t* could expect to infect given that conditions remain unchanged (Fraser, 2007). Let *I*(*t*) be the total number of infections by viruses of any genotype of *a* or *A*_1_, …, *A*_*n*_ at calendar time *t* and *g*(*j*) be the probability mass function of serial intervals. Suppose that *g*(*j*) is small enough to be neglected for *j* < 1 and *j* > *l*. The instantaneous reproduction numbers of genotypes *a* and *A*_1_, …, *A*_*n*_ at calendar time *t* are represented as follows:

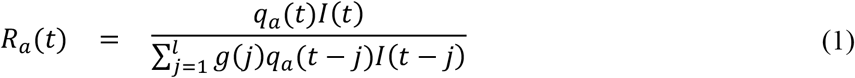

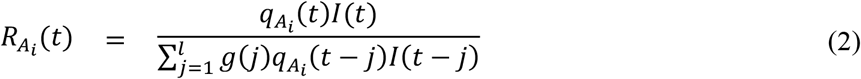

Since a virus of genotype *A*_*i*_ generates 1 + *s*_*i*_ times as many secondary transmissions as those of genotype *a*, the following equation holds

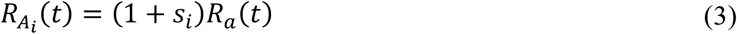

for each calendar time *t* ≥ *t*_*i*_. We call the value of 1 + *s*_*i*_ the relative instantaneous reproduction number of *A*_*i*_ with respect to *a*.

Next, we assume that for all infections at calendar time *t*, the difference in the number of infections at the time when previous generations infected can be regarded as considerably small, i.e.

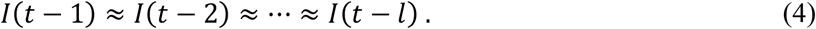

The frequency of genotype *A*_*i*_ in the viral population at calendar time *t*, 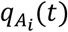, can be modeled as follows:

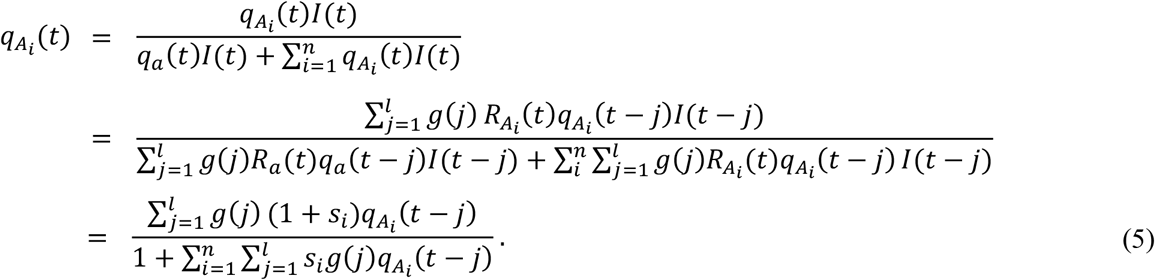

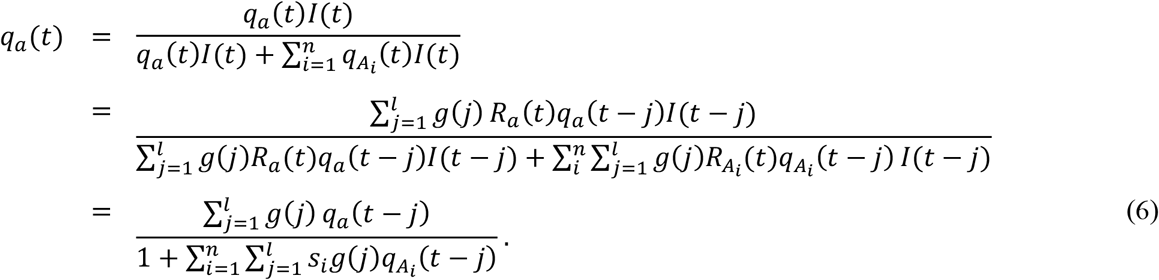

## Numbers of SARS-CoV-2 variants detected in Japan

Metadata of nucleotide sequences of SARS-CoV-2 submitted from Japan since December 1, 2020 were downloaded from the GISAID EpiCoV database (Shu & McCauley, 2017) on June 16, 2021. After removing sequence records of viruses detected at Airport Quarantine Station in Japan, PANGO lineage labels (Rambaut et al., 2020) assigned to these sequences were collected (Supplementary Table 1). Numbers of sequences assigned to the R.1, Alpha (B.1.1.7), or Delta (B.1.617.2) were counted and those of other lineages were summed up to “other” linages. Weekly numbers of the R.1 (E484K), Alpha (N501Y), and Delta (L452R) variants detected by PCR in Tokyo (Supplementary Table 3) were obtained from the report on COVID-19 monitoring submitted by Tokyo Metropolitan Government on June 17, 2021 (Tokyo Metropolitan Government, 2021). The GISAID variant counts after April 26, 2021 and The PCR detection counts before April 26, 2021 were excluded from the analysis to avoid double counting. Finally, daily variant frequencies of a total of 28,211 sequences—consisting of 5,186 R.1, 8,205 Alpha, 2 Delta, and 14,818 other viruses—and weekly variant frequencies of 784 viruses—consisting of 88 R.1, 664 Alpha, and 32 Delta were obtained and used in the rest of analyses (Supplementary Table 2; Supplementary Table 3).

## Relative instantaneous reproduction numbers

Let *N*(*t*) be the number of sequences of either genotype *A*_1_, …, *A*_*n*_, or *a* observed at calendar time *t*, and let *d*_1_, …, *d*_*k*_ be calendar times such that *N*(*d*_*j*_) > 0 for 1 ≤ *j* ≤ *k*. Suppose that we have 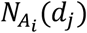 sequences of genotype *A*_*i*_ at calendar time *d*_*j*_ for 1 ≤ *i* ≤ *n* and 1 ≤ *j* ≤ *k*. Since genotype *A*_*i*_ emerged at time 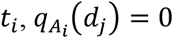 for *d*_*j*_ < *t*_*i*_. If the genotype *A*_*i*_ emerges with an initial frequency of *q*_*i*_(*t*_*i*_) at calendar time *t*_*i*_, then the likelihood function of parameters *s*_1_, …, *s*_*n*_ and *q*_1_(*t*_1_), …, *q*_*n*_(*t*_*n*_) for observing 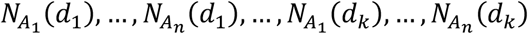 sequences of genotype *A*_1_, …, *A*_*n*_, at calendar times *d*_1_, …, *d*_*k*_ is given by the following formula:

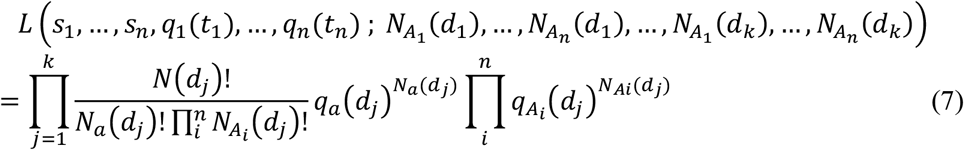

where 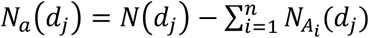 for 1 ≤ *j* ≤ *k*.

The earliest dates of the R.1, Alpha, and Delta variants in the GISAID sequence from Japan (excluding those from Airport Quarantine Station) were December 1, 2020, December 15, 2020, and April 15, 2021, respectively. We assume that *t*_*R*.1_, *t*_*Alpha*_, and *t*_*Delta*_ are these dates. The serial intervals were assumed to follow a lognormal distribution with *μ* = 1.38 and *σ* = 0.563 discretized and truncated so that *g*(0) = 0 and *g*(*j*) = 0 for *j* > 0. Parameters *s*_*R*.1_, *s*_*Alpha*_, *s*_*Delta*_, *q*_*R*.1_, *q*_*Alpha*_, and *q*_*Delta*_were estimated by maximizing the likelihood defined in Equation (7). The 95% confidence intervals (CIs) of parameters *s*_*R*.1_, *s*_*Alpha*_, *s*_*Delta*_, *q*_*R*.1_(*t*_*R*.1_), *q*_*Alpha*_(*t*_*Alpha*_), and *q*_*Delta*_(*t*_*Delta*_) were estimated by profile likelihood (Pawitan, 2013). The optimization of likelihood function and calculation of 95% confidence intervals were performed using the nloptr package in R (Johnson, 2020; Rowan, 1990).

The selective advantages *s*_*R*.1_, *s*_*Alpha*_, and *s*_*Delta*_ with respect to other strains circulating in Japan were estimated to be 0.245 (95% CI: 0.245–0.246), 0.437 (95% CI: 0.435–0.440), and 0.948 (95% CI: 0.896–0.993), assuming that the serial interval distribution is a lognormal distribution with a log mean of 1.38 and log standard deviation of 0.563 (Table 1). The initial frequencies, *q*_*R*.1_(*t*_*R*.1_), *q*_*Alpha*_(*t*_*Alpha*_), and *q*_*Delta*_(*t*_*Delta*_), were estimated to be 0.00430 (95% CI: 0.00411– 0.00455), 0.00114 (95% CI: 0.00103–0.00115), and 0.00290 (95% CI: 0.00228–0.00331), respectively (Table 1). The relative instantaneous reproduction number of the Delta was consistent with the result using global data by Cambell et al., although a considerable difference was observed for the Alpha (Campbell et al., 2021).

**Table 1.**
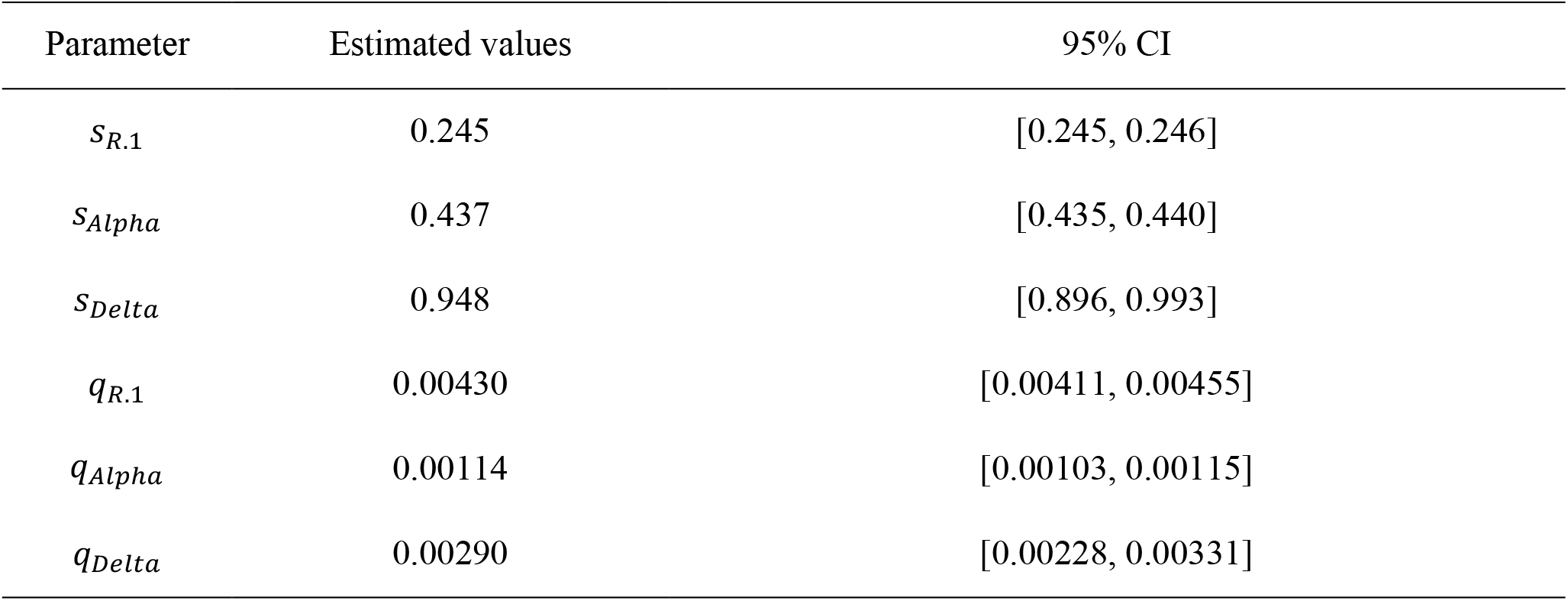
Estimated values of parameters and their 95% CIs.

Figure 1 shows temporal changes in frequencies of R.1, Alpha, Delta variants, and other strains circulating in Japan estimated by our model. As can be observed from the, the Alpha seems to be predominant in Japan at the beginning of June 2021. However, the Delta is predicted to take over the Alpha on July 12, 2021 with a 95% CI from July 5 to July 22, 2021.

**Figure 1.**
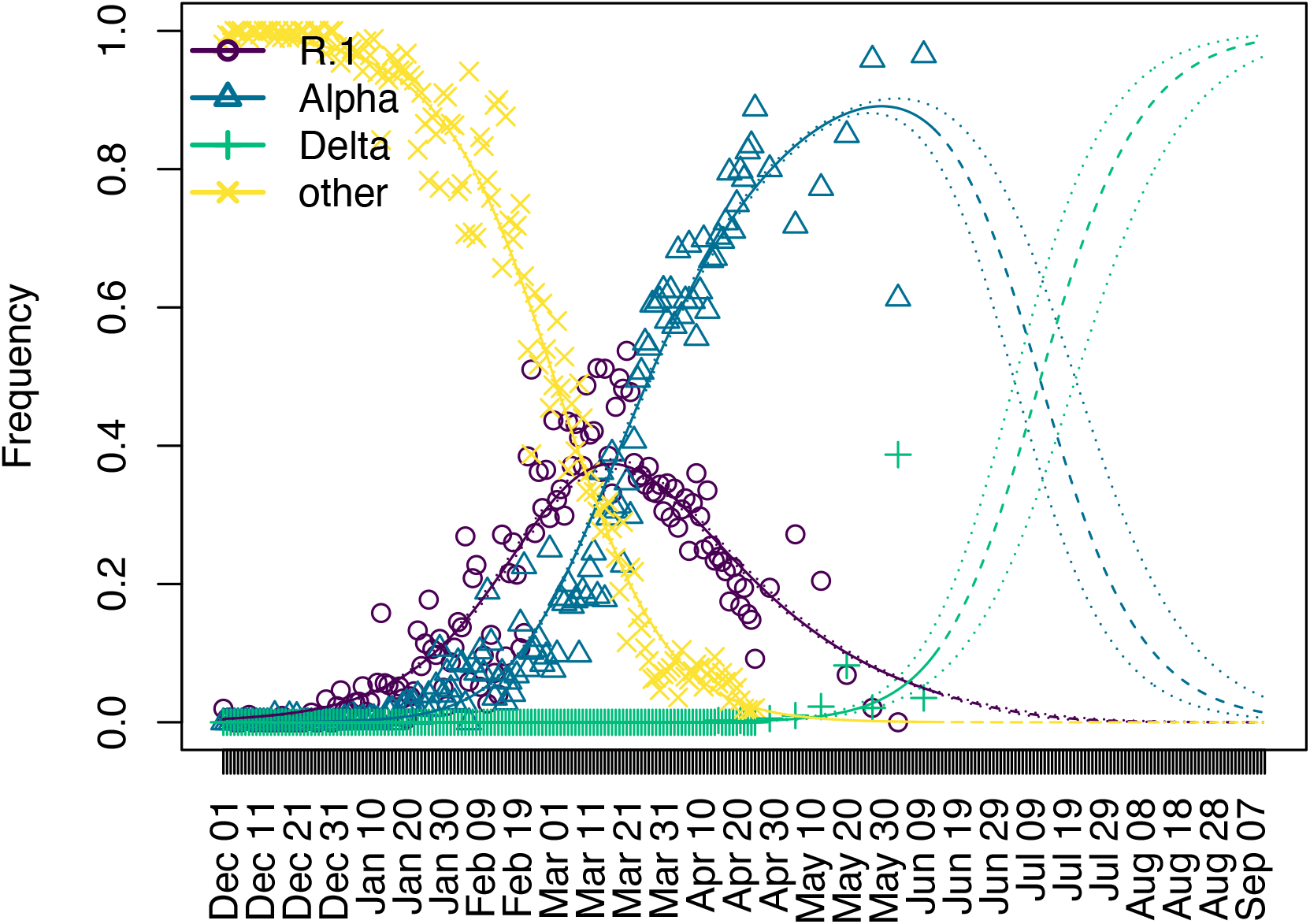
Temporal changes in variant frequencies of R.1 (purple), Alpha (blue), Delta (green), and others (yellow) circulating in Japan from December 1, 2020 to September 11, 2021. The circles, triangles, pluses, and crosses respectively indicate frequencies of SARS-CoV-2 sequences of R.1, Alpha, Delta, and others collected in Japan from December 1 2020. Daily frequencies are shown until to April 25, 2021 and weekly frequencies from April 26. The solid lines indicate the maximum likelihood estimates of frequencies of the variants using collected data. The dashed lines represent the predicted variant frequencies after June 13, 2021. Dotted lines indicate lower and upper bounds of 95% CI of estimated variant frequencies.

## Population average of relative instantaneous reproduction numbers

The population average of relative instantaneous reproduction numbers, 1 + *s*, with respect to strains labeled as others at calendar time *t* was estimated by 1 + *s*_*R*.1_*q*_*R*.1_(*t*) + *s*_*Alpha*_*q*_*Alpha*_(*t*) + *s*_*Delta*_*q*_*Delta*_(*t*). Since the maximum likelihood estimates of *s*_*R*.1_, *s*_*R*.1_, and *s*_*Delta*_ can be affected by the log mean and log standard deviation of the lognormal serial interval distribution, sensitivity analyses of parameters *s*_*R*.1_, *s*_*Alpha*_, *s*_*Delta*_ were performed by using the combinations of *μ* and *σ* sampled along the boundary of the 95% confidence area of likelihood surface of the serial interval distribution (Nishiura, Linton, & Akhmetzhanov, 2020 ; Piantham et al., 2021). As a result, we estimated the ranges of the maximum likelihood estimates of *s*_*R*.1_, *s*_*R*.1_, and *s*_*Delta*_ to be 0.190–0.319, 0.335–0.580, and 0.703–1.30, respectively.

Figure 2 shows temporal changes in the population average of relative instantaneous reproduction numbers with respect to other strains than the R.1, Alpha, or Delta circulating in Japan. The population average of relative instantaneous reproduction numbers stayed around one until the end of January 2021, since the strains other than the R.1, Alpha, or Delta variants were predominant around that time. From the beginning of February 2021, the population average of relative instantaneous reproduction has been elevated due to the increase of frequencies of the Alpha variant (Figure 1). The population average of relative instantaneous reproduction numbers reached at 1.20 on March 9, 2021. It should be noted that around this day the decrease in the number of COVID-19 cases in Tokyo increased although Tokyo was under the declaration of emergency status. From around the middle of June 2021, the population average of relative instantaneous reproduction numbers elevates because the frequencies of the Alpha started to decrease due to the replacement of the Alpha by the Delta (Figure 1). The elevation of population average of relative instantaneous reproduction numbers will continue until the Alpha is completely replaced by the Delta the middle of August 2021 (Figure 2).

**Figure 2.**
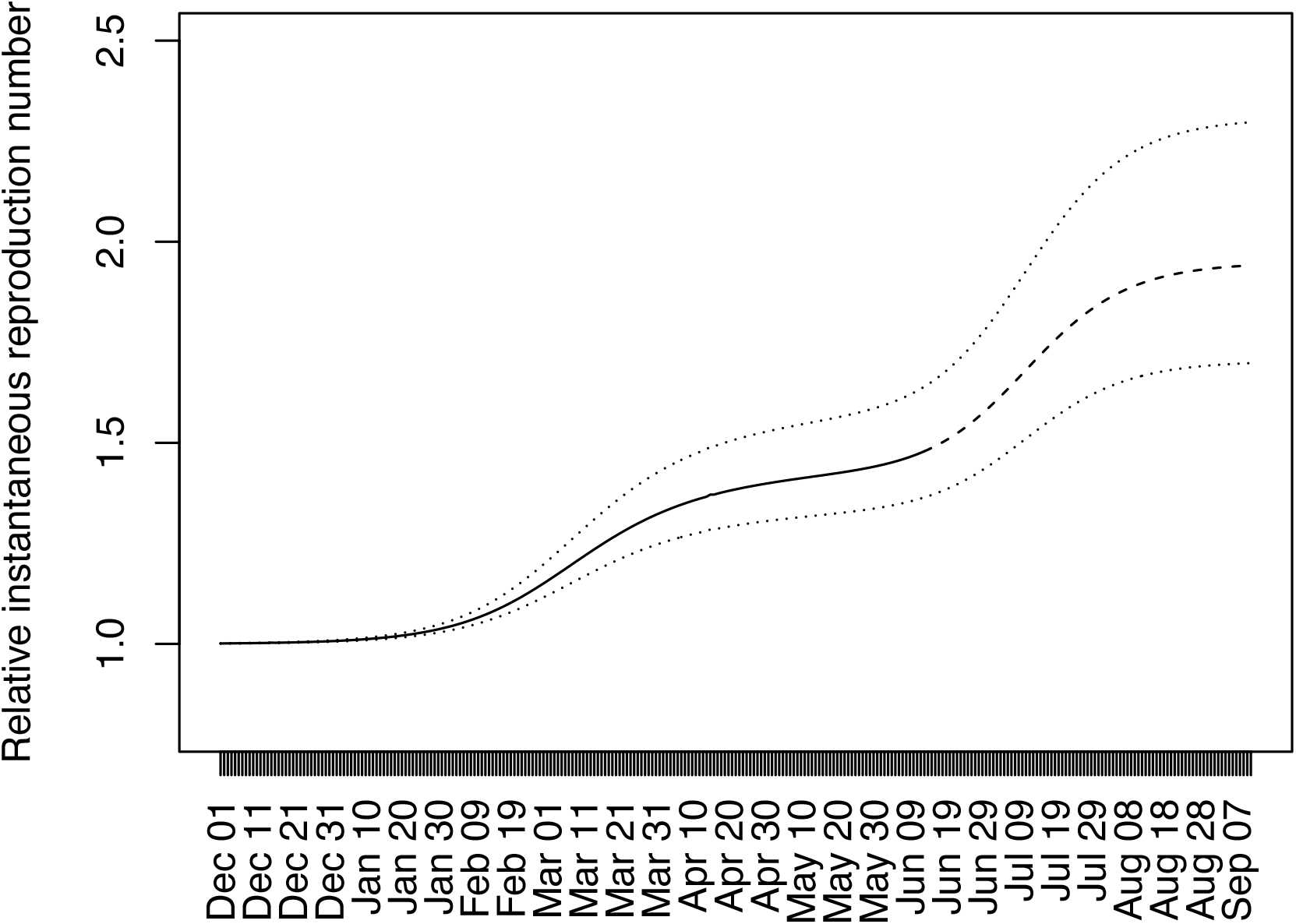
Temporal changes in the population average of the relative instantaneous reproduction numbers with respect to strains other than the R.1, Alpha, or Delta in Japan. The solid line (until June 13, 2021) and dashed line (from June 14, 2021) indicate the population average of the relative instantaneous reproduction numbers when *s*_*R*.1_ = 0. 245, *s*_*Alpha*_ = 0. 437, and *s*_*Delta*_ = 0. 948, which are calculated by assuming the lognormal serial interval distribution with *μ* = 1. 38 and *σ* = 0. 563. The lower dotted line indicates the population average of the relative instantaneous reproduction numbers when *s*_*R*.1_ = 0. 190, *s*_*Alpha*_ = 0. 335 and *s*_*Delta*_ = 0. 703, which are the lower bounds of *s*_*R*.1_, *s*_*Alpha*_, and *s*_*Delta*_ calculated based of the 95% confidence area of the lognormal serial interval distribution. The upper dotted line indicates those when *s*_*R*.1_ = 0. 319, *s*_*Alpha*_ = 0. 580 and *s*_*Delta*_ = 1. 30, which are upper bounds calculated in the same way.

### Tokyo Olympic games

We have shown that the Delta variant possesses evidently greater transmissibility than the Alpha variant. The relative instantaneous reproduction number of the Alpha with respect to other strains circulating in Japan was estimated at 1.437 and those of the Delta was estimated at 1.948. This means that the Delta possesses 1.356 times higher transmissibility than the Alpha. While the Alpha variant has just replaced other variants in Japan over the last 5 months, it is very likely that it is just a matter of time for the Delta variant to replace others including the Alpha.

An important learning point of this rapid communications is that, unfortunately, the replacement is likely to happen mostly before the Tokyo Olympic games from July 23, 2021. The risk assessment must account for the fact that substantial number of international visitors during the Games will be exposed to the Delta variant, and an increased mobility could help further spread COVID-19 caused by this variant with an elevated transmissibility around the world.

Already we have seen that the interventions have had to be strengthened with the Alpha variant. We have just shown that the focused interventions on drinking and eating services that have been highly effective over the last one year was not effective to substantially reduce the reproduction number below the value of one. However, the Delta variant may require even more.

## Data Availability

The raw data used for the analysis were provided as Supplementary Table 1.

## Acknowledgement

We gratefully acknowledge the laboratories responsible for obtaining the specimens and the laboratories where genetic sequence data were generated and shared via the GISAID Initiative, on which this research is based. The information on originating laboratories, submitting laboratories, and authors of SARS-CoV-2 sequence data can be found in Supplementary Table 2. This work was supported by the Japan Agency for Medical Research and Development (grant numbers JP20fk0108535). K.I. received funding JSPS KAKENHI (21H03490). P.C. was supported by the World-leading Innovative and Smart Education Program (1801) from the Ministry of Education, Culture, Sports, Science, and Technology, Japan. H.N. received funding from Health and Labor Sciences Research Grants (20CA2024 and 20HA2007); the Japan Agency for Medical Research and Development (JP20fk0108140); JSPS KAKENHI (21H03198) and the Japan Science and Technology Agency (JST) SICORP (e-ASIA) program (JPMJSC20U3). The funders had no role in the study design, data collection and analysis, decision to publish, or preparation of the manuscript.

